# Sleep increases broadband fast oscillations in the epileptic focus of patients with focal cortical dysplasia

**DOI:** 10.1101/2025.02.18.25322331

**Authors:** Nina Schwoon, Mohammad F Khazali, Armin Brandt, Matthias Dümpelmann, Yiwen Li Hegner, Nicolas Roehri, Dirk-Matthias Altenmüller, Victoria San Antonio-Arce, Peter C Reinacher, Julia M Nakagawa, Soroush Doostkam, Theo Demerath, Horst Urbach, Andreas Schulze-Bonhage, Marcel Heers

## Abstract

**Rationale:** Focal cortical dysplasia (FCD) is associated with increased seizure-risk during sleep, yet the underlying mechanisms remain unclear. Previous studies report increased oscillatory patterns during sleep in focal epilepsy patients with FCD. We investigated whether fast oscillations (FOs, 14-250 Hz) within the epileptic focus significantly change during sleep compared to wake, hypothesizing that analyzing broad frequency bands provides more comprehensive information than narrow bands alone.

**Methods:** We retrospectively analyzed intracranial EEG recordings from 22 FCD patients (2010-2023), focusing on contacts within the irritative zone (IZ) and seizure onset zone (SOZ). Using semiautomated detections, we compared FO rates between one-hour wake and sleep epochs across beta (14-40 Hz), gamma (40-80 Hz), and ripple (80-250 Hz) bands. Distance-based Multivariate Analysis of Variance (MANOVA) was applied to integrate the spatial organization of FO changes, enabling simultaneous analysis of individual and combined broadband ranges (BGR) while accounting for neocortical detection rates outside the epileptic focus and eloquent cortex.

**Results:** Analysis of 67 ± 28 (median ± standard deviation) bipolar iEEG contact pairs per patient (SOZ: 14 ± 12, IZ: 29 ± 16) revealed distinct sleep-related patterns. In the SOZ, gamma showed significant increases in 10 patients, followed by BGR in seven patients. The IZ exhibited strong changes in both BGR and gamma (13 patients each) with high concordance (12 matches). Beta and ripple bands showed moderate concordance. Notably, gamma oscillation rates in the SOZ specifically increased in patients with sleep-related epilepsy (p < 0.05).

**Conclusion:** Gamma oscillations showed the most robust sleep-related increases in the SOZ, while both gamma and BGR showed strong changes in the IZ. These findings suggest gamma oscillations, complemented by broadband analysis, may serve as reliable markers for sleep-related changes in FCD patients.

**Funding:** YLH (LI1904/2-1) and MH (HE6844/3-1; project ID for both: 468174690) were funded by the German Research Foundation (DFG)

**Key points:**

- Within the seizure onset zone, gamma band oscillations (40-80 Hz) followed by combined broadband BGR (14-250 Hz) show more frequent significant increases from wake to sleep across patients
- Within the irritative zone, gamma band and broadband oscillations BRG show even more frequent increases from wake to sleep across patients
- Changes occur simultaneously across multiple frequency bands, supporting the value of multi-band analysis
- Gamma oscillation rates within the seizure onset zone significantly increased from wake to sleep in patients with sleep-related epilepsy (p < 0.05)

## Introduction

### Epilepsy

Epilepsy is one of the most common chronic neurological diseases (World Health Organization, 2019). Approximately 30% of patients with focal epilepsy suffer from drug resistance (Schuele and Lüders, 2008), defined as the absence of sustained seizure freedom after adequate therapy with at least two antiseizure medications (ASMs) (Kwan *et al*., 2010). Epilepsy surgery can be a viable therapeutic option in these patients, achieving seizure freedom in 60-65 % of patients after two years, particularly for patients with lesions visible in the MRI (Wagstyl *et al*., 2022). Pre-surgical diagnostics determines the characteristics of the epileptic focus and the feasibility of epilepsy surgery. It contains high-resolution MRI of the brain, neuropsychological testing and non-invasive video-EEG monitoring to identify the irritative zone (IZ) where interictal epileptiform discharges occur and the seizure onset zone (SOZ), where seizures arise from (Rosenow and Lüders, 2001). In a subset of patients, invasive video-EEG monitoring with intracranial EEG (iEEG) needs to be added to delineate the SOZ and the IZ at a higher spatial resolution (Rosenow and Lüders, 2001). Among the pathologies requiring such detailed pre-surgical evaluation, focal cortical dysplasia (FCD) represents one of the most frequent causes of drug-resistant epilepsy.

### Focal cortical dysplasia and sleep-related epilepsy

Understanding the pathophysiology of FCD and its relationship to sleep is crucial for improving therapeutic strategies in drug-resistant epilepsy (Nobili *et al*., 2009; Chassoux *et al*., 2012; Wang *et al*., 2022). Five to twenty-five percent of patients with focal epilepsy suffer from FCD (Palmini *et al*., 1995; Bast *et al*., 2006). FCDs are cortical developmental disorders caused by a malfunction in neuronal migration and differentiation (Hildebrandt and Blümcke, 2004). Notably, FCD distinctly increases the risk of sleep-related epilepsy (SRE) (Wang *et al*., 2022), where the majority of seizures occur during sleep (Nobili *et al*., 2009). Among the three subtypes of focal cortical dysplasia (FCD), type II is distinguished by the presence of dysmorphic neurons and, in the case of type IIB, balloon cells (Palmini *et al*., 2004; Blümcke *et al*., 2011). Notably, FCD type II is also associated with an increased risk of sleep-related epilepsy (SRE) (Nobili *et al*., 2009).

### Differences of fast oscillations (FOs) between wake and sleep

The relationship between FCD and sleep-related seizures is observed in intracranial EEG (iEEG) recordings, by the presence of characteristic patterns. These patterns span a broad range of frequencies from 14-250 Hz (Tassi, 2002; Heers *et al*., 2018) and differ markedly between wakefulness and sleep (Chassoux *et al*., 2012). Visual analysis reveals that repetitive sharp waves predominate during wakefulness, whereas rhythmic patterns in different frequency bands are more prevalent during sleep (Menezes Cordeiro *et al*., 2015; Eltze *et al*., 2020). Semi-automatic detection methods can identify these rhythmic patterns, known as FOs, in specific frequency bands: beta (14-40 Hz), gamma (40-80 Hz), and ripple (80-250 Hz) (Roehri *et al*., 2016, 2017).

Different frequency bands reflect distinct yet potentially overlapping aspects of FCD pathophysiology. While these bands have been individually studied in relation to the epileptic focus (Aubert *et al*., 2009; Sato *et al*., 2017; Heers *et al*., 2018), their complex interplay during wake and sleep states remains poorly understood. FOs above 80 Hz spatially correlate with the epileptic focus and reflect disease severity in FCD patients (Kerber *et al*., 2013) and they show increased expression during sleep compared to wakefulness (Frauscher *et al*., 2015).

### Hypothesis

We hypothesized that FO rates across a broad frequency range (14-250 Hz) within the SOZ and the IZ would show systematic differences between non-rapid eye movements (NREM) sleep stages 2-3 and wakefulness. Therefore, a holistic analysis of the spatial correlations of broadband frequencies considering beta, gamma and ripple oscillations would be more informative about sleep-related changes of the epileptic focus consisting of the IZ and the SOZ than focusing on narrow frequency bands alone.

## Patients and methods

### Patient selection

All 22 patients underwent iEEG at the Epilepsy Center of the University Hospital Freiburg between 2009 and 2023. Initially, 31 patients above the age of 12 years with suspected neocortical FCDs were reviewed. Seven patients were excluded due to a lack of continuous iEEG data, and two patients were excluded because seizures were too frequent to select a sufficient interictal time interval for analysis. The mean age of patients was 25.3 ± 12.8 (mean ± standard deviation) years with a mean age at first manifestation of 11.6 years (STD ± 10.5) and a disease duration of 17.7 ± 12.3 years. Twelve patients were female (more details on included patients in Table 1, details on iEEG, histopathology and postsurgical outcomes in Suppl. 1).

**Table 1:**
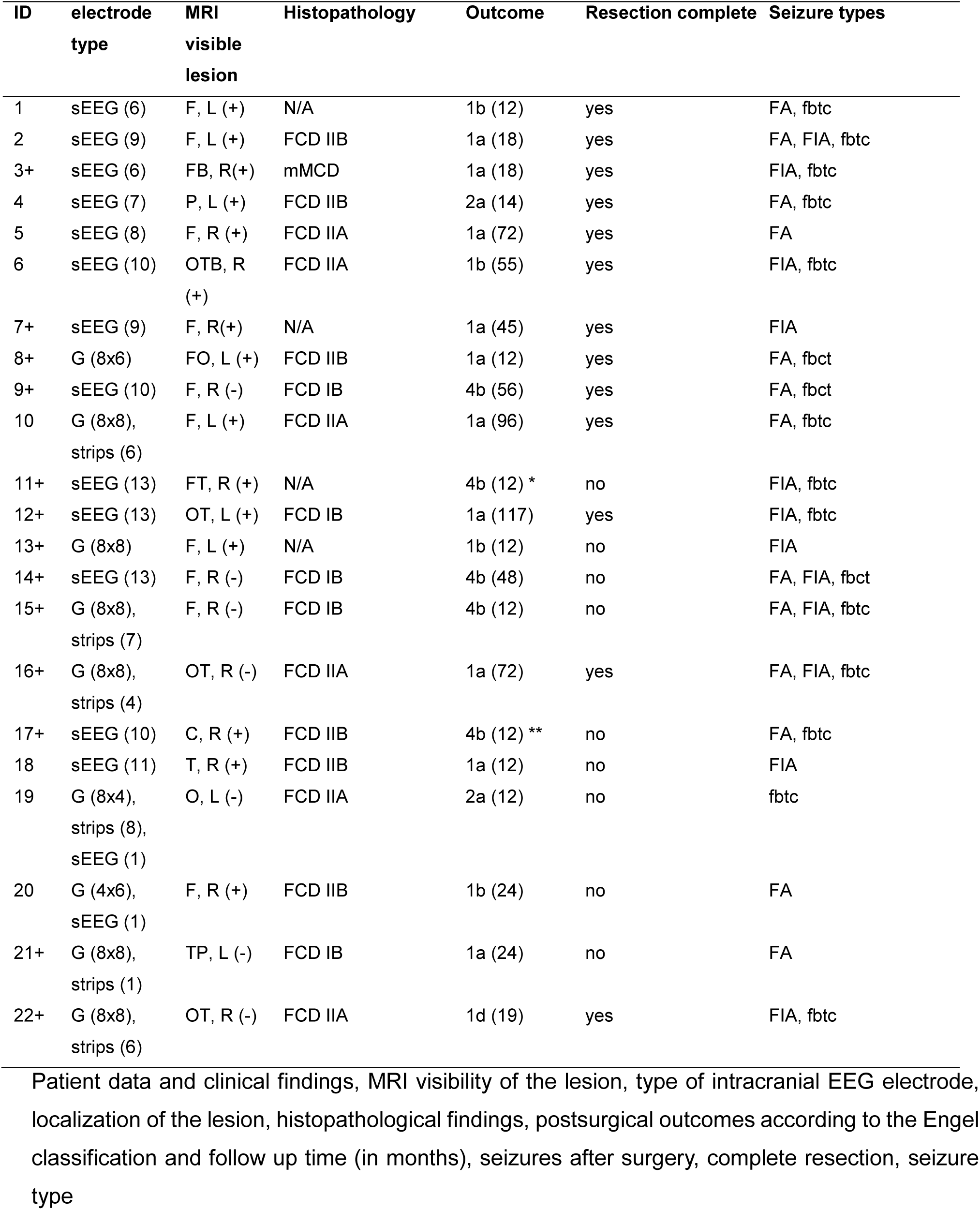
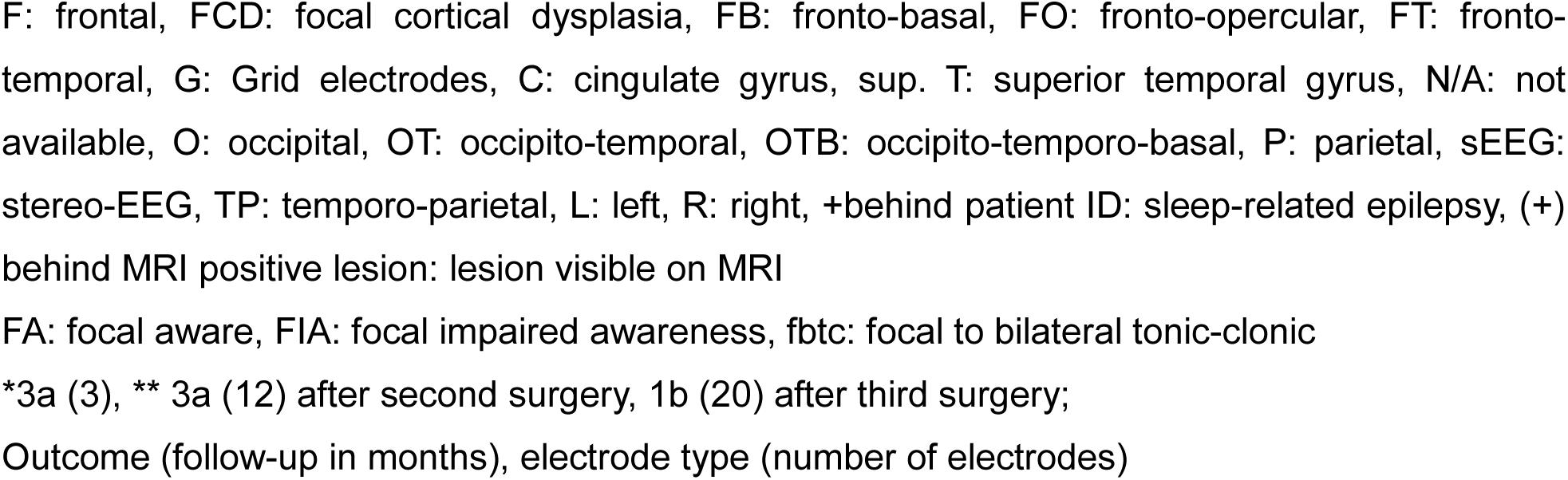
Clinical characteristics of included patients.

### iEEG analysis

For each patient, one interval during sleep and one interval during wakefulness were selected. These intervals each had a duration of 1 hour, ended at least 15 minutes before and started at least 30 minutes after focal aware or focal impaired awareness seizures. After focal to bilateral tonic-clonic seizures, a minimum time interval of 2 hours was required before the selected interval. For sleep analysis, we aimed to select intervals predominantly consisting of NREM sleep of stages 2 or 3 according to the criteria of the American Society of Sleep Medicine and Rechtschaffen and Kahles (Moser *et al*., 2009). For patients with available surface EEG data (n=11 patients), Persyst 14c (Persyst Inc., Solana Beach, CA, USA) was used for initial sleep stage screening. Based on Persyst screening results, sleep intervals consisting predominantly of stages NREM2 or NREM3 were selected. When surface EEG was unavailable (n=11 patients), sleep and wake intervals were determined by an experienced and certified epileptologist (M.H.) through visual evaluation of video monitoring and iEEG data.

The iEEG data were recorded using Neuvo amplifiers (Compumedics Neuroscan, Abbotsforth, Victoria, Australia). All recordings had a sampling rate of 2 kHz except for patient ID 11, where only iEEG data with a sampling rate of 1 kHz were available. The iEEG data were visually reviewed in multiple montages with AnyWave software (Colombet *et al*., 2015), using a high-pass filter with a cutoff frequency of 0.3 Hz, without low-pass filtering. EEG channels with muscle artifacts due to an inverse breach rhythm (Fiederer *et al*., 2016) or technical artifacts were removed from the analysis. Within AnyWave, the Delphos toolbox (Roehri *et al*., 2018) was used to detect FOs in the beta (12-40 Hz), gamma (40-80 Hz), and ripple (80-250 Hz) bands using a bipolar montage (Figure 1).

**Figure 1:**
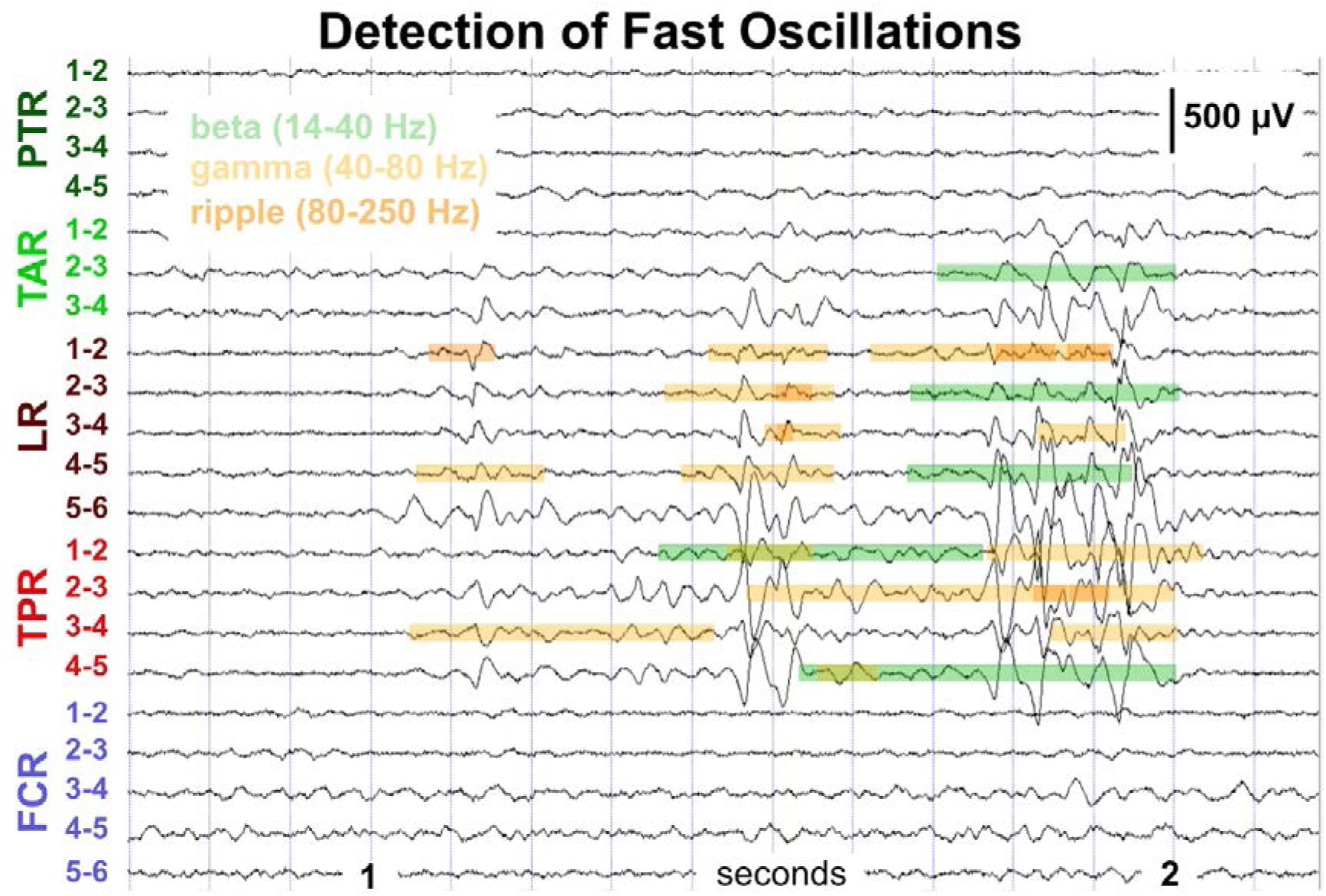
Detection of fast oscillations (FOs) using the toolbox Delphos (Roehri *et al*., 2016) within the AnyWave software (Colombet *et al*., 2015). Signal traces of bipolar channels of five intracranial Stereo-EEG electrodes shown (high-pass filter: 1 Hz, notch filter: 50 Hz, no low-pass filter). Markers for beta (14-40 Hz, green), gamma (40-80 Hz, yellow) oscillations and ripples (80-250 Hz, orange) shown on marked EEG traces. Note the spatiotemporal overlap of different types of markers reflecting the co-occurrence of FOs in different frequency bands of intracranial EEG recordings. Illustration of P12 (Table 1), same patient as Figure 2.

### Classification of iEEG contacts

Based on the available clinical data, iEEG contacts were classified according to Lüders’ model of the organization of the epileptic focus (Lüders, Burgess and Noachtar, 1993; Lüders *et al*., 2006). The iEEG contacts for each patient were categorized into two regions: the seizure onset zone (SOZ) containing 14 ± 12 contacts, and an irritative zone (IZ) containing 29 ± 16 contacts. In rare cases where SOZ channels were not part of the clinical IZ definition, they were included in the IZ group for this evaluation. Contacts within the eloquent cortex regions were marked as ELOQ. To determine electrodes implanted in eloquent brain areas, the results of clinical electrical stimulation were used. In patients without electrical stimulation, electrodes within the eloquent cortex were identified using the Automatic Anatomical Labeling atlas (Tzourio-Mazoyer *et al*., 2002). Only electrodes located in eloquent cortex regions with a probability of > 70% according to the atlas were included in the ELOQ group.

Contacts outside the mesial temporal lobe, the IZ, the SOZ and the ELOQ group were classified as OTHER (30 contacts ± 21) resulting in a group of neocortical electrode contacts to adjust for widespread neocortical sleep-related changes in FO rates. As we evaluated bipolar iEEG signals, iEEG electrode contacts were classified as being part of the SOZ or the IZ if at least one of the two contacts of the bipolar montage was part of the SOZ or IZ. If there was an overlap between the ELOQ group and the SOZ or IZ, these contacts were classified as SOZ or IZ contacts.

### Wake and sleep effect on FO rates in different frequencies

To identify FO rates that were specific for the epileptic focus (SOZ or IZ), we subtracted the median FO rates of electrode contact pairs in the group OTHER from the rates of electrode contact pairs within the epileptic focus. The FO rates were evaluated for the SOZ and IZ for each frequency band (beta, gamma, and ripple) separately for wake and sleep states (example patient, Figure 2). To evaluate the effect of the brain states during wake and sleep on FOs in each frequency band, we tested whether FO rates increased from wake to sleep, separately for each frequency band, using non-parametric Wilcoxon rank sum tests for median FO rates across all patients (Figure 3). For the patient-by-patient analysis, we employed distance-based Multivariate Analysis of Variance (MANOVA)(Anderson, 2001; Jones, 2017). This method offers the following advantages: 1: MANOVA calculates the distance of changes in FO rates while accounting for the spatial organization of detection rates through a Euclidean distance matrix across all contacts. 2: Simultaneous multiband analysis. The method allows for the inclusion of FO rates from all bands – beta, gamma, and ripple (BGR) – simultaneously. Each band’s FO rate is represented on an axis within a 3D feature space, creating one data point for wake and one for sleep (Figure 4A-D). The Euclidean distance between the sleep and wake states in this 3D space is then calculated to perform MANOVA. We applied MANOVA to analyze FO rates for all bands combined (BGR) as well as for each band individually, ensuring a fair comparison between the combined BGR and the individual bands (Figure 4). Using MANOVA analysis allows processing the same data sample size independently of the number of bands included in the analysis. For a single band, the data are represented in a 1D space, whereas for BGR, the data are represented in a 3D space. In all cases, the sample size is determined by the number of contacts, which remains constant for a given patient. This approach provides a robust framework for analyzing FO rate changes across wake and sleep states, capturing the multidimensional complexity of BGR dynamics. While analyzing more frequency bands typically improves our ability to distinguish between sleep and wake states, this advantage diminishes when different bands show conflicting patterns of change.

**Figure 2:**
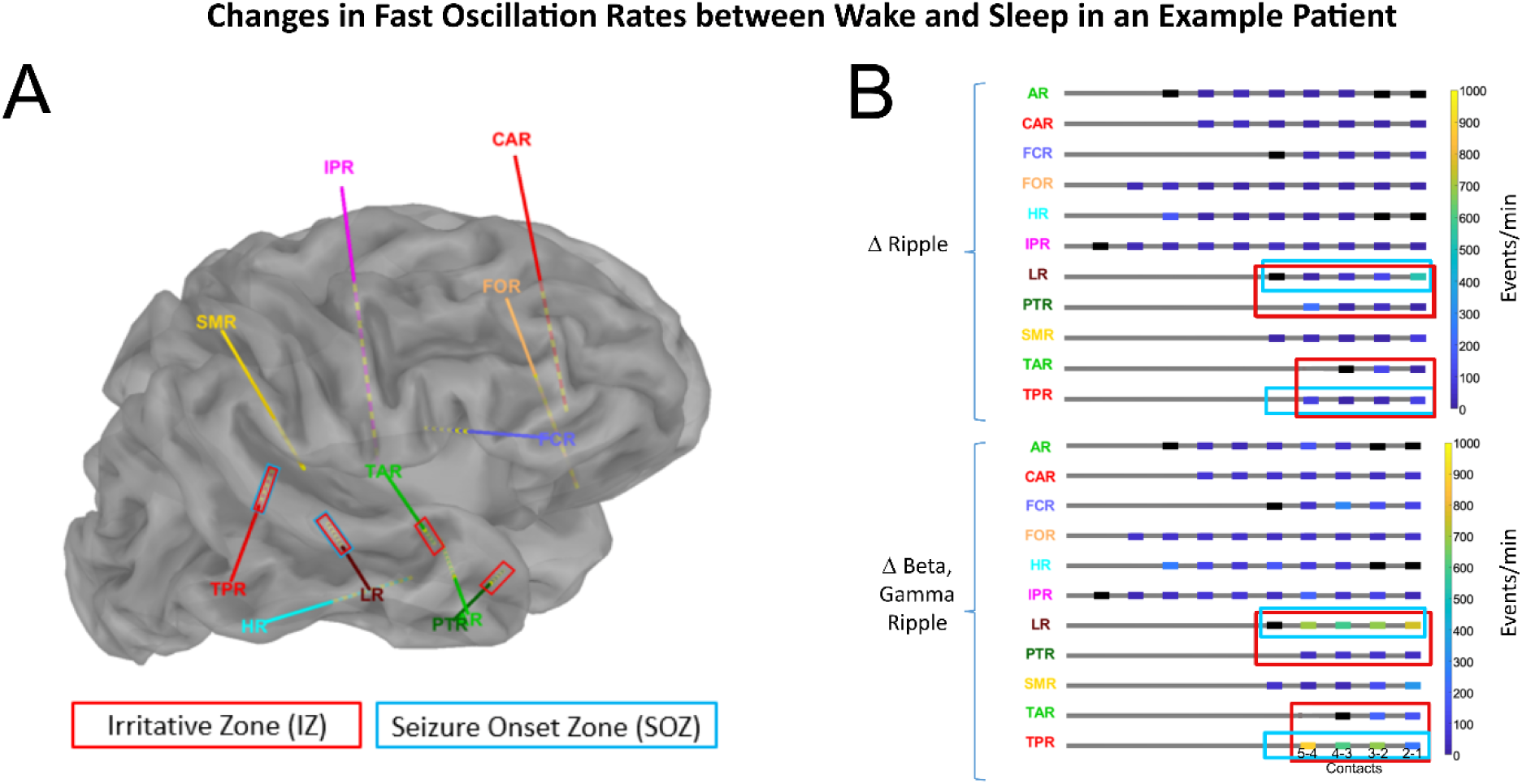
Detection of fast oscillations in an example patient with a focal cortical dysplasia type II in the right posterior superior temporal gyrus. A: Implantation scheme with nine invasive Stereo-EEG (iEEG) electrodes implanted in the right frontal, central, parietal, insular and temporal cortex. The irritative zone (IZ) contacts are marked with red rectangles and the seizure onset zone (SOZ) contacts are marked with blue rectangles. B: Rate increase of ripples (upper part) and of broadband BGR (lower part) for each iEEG electrode from wake to sleep iEEG recordings. Please note the more pronounced detection rate changes when considering rate changes in the beta, gamma, and ripple frequency band together (lower panel) instead of focusing on the ripple frequency band alone (upper panel). Illustration of P12 (Table 1), same patient as Figure 1.

**Figure 3:**
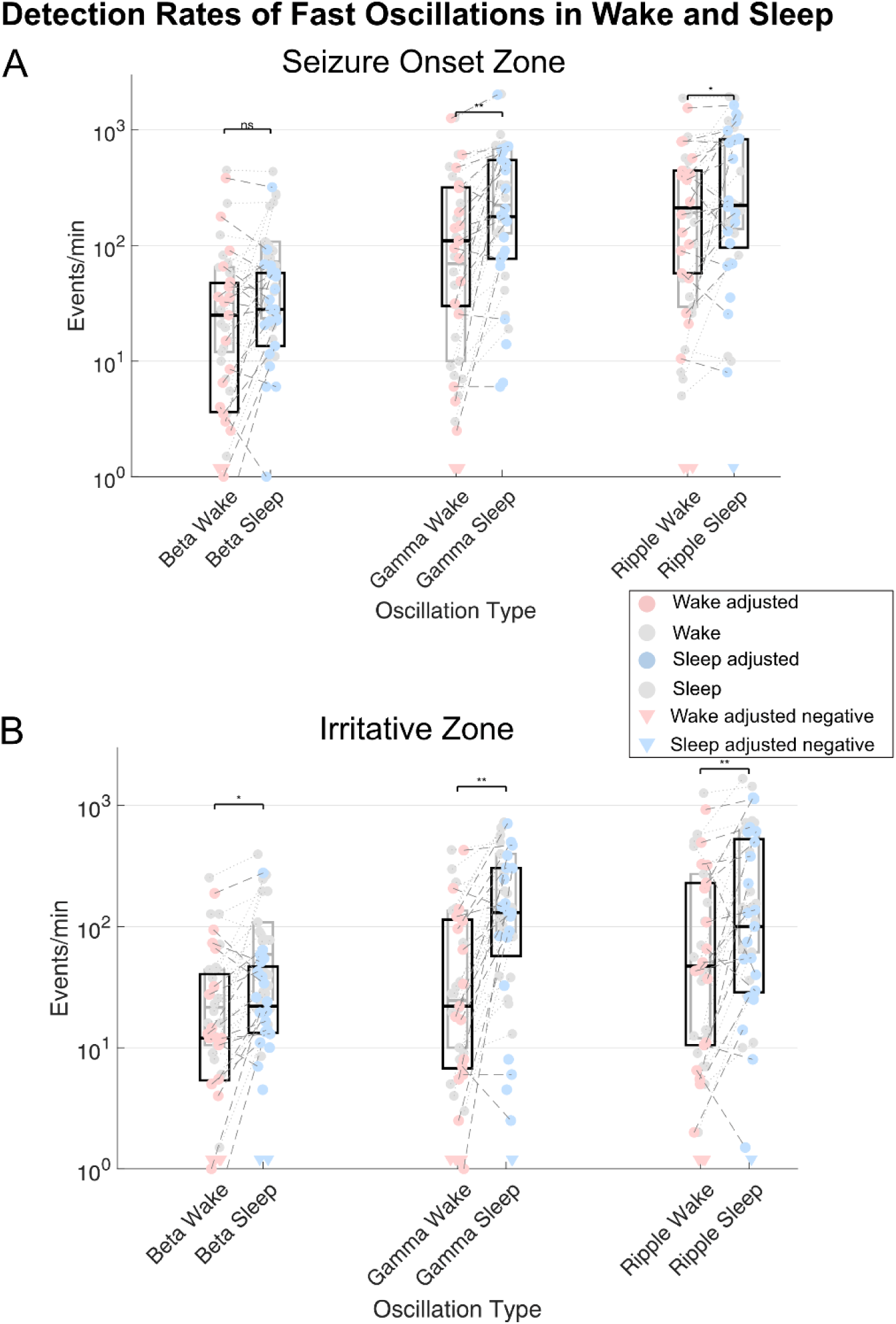
Comparison of detection rates between wake and sleep for beta and gamma oscillations and ripples across all patients. A: presents the rates in the seizure onset zone (SOZ). B: presents the rates in the irritative zone (IZ). Raw detection rates within the SOZ or IZ are illustrated as raw dots connected with grey lines between wake and sleep. Detection rates adjusted by subtracting the oscillations rates measured in the electrodes group OTHER (details in methods: Classification of iEEG contacts) are illustrated in magenta (wake) and blue (sleep). Boxplots for raw detections are shown in gray and boxplots for adjusted detections in black. Significance level: ** p < 0.01, *: p < 0.05, ns: not significant. Note that for the SOZ we a see substantial increase for the adjusted gamma oscillations and ripples between wake and sleep (FDR-corrected Wilcoxon-test), but not for the beta frequency band. For the IZ we see a substantial increase for the adjusted beta and gamma oscillations and ripples (FDR-corrected Wilcoxon-test). Note that the gray and black boxes of ripples in SOZ are largely overlapping with no correction effect, which is likely because ripples are very specific to the SOZ.

**Figure 4:**
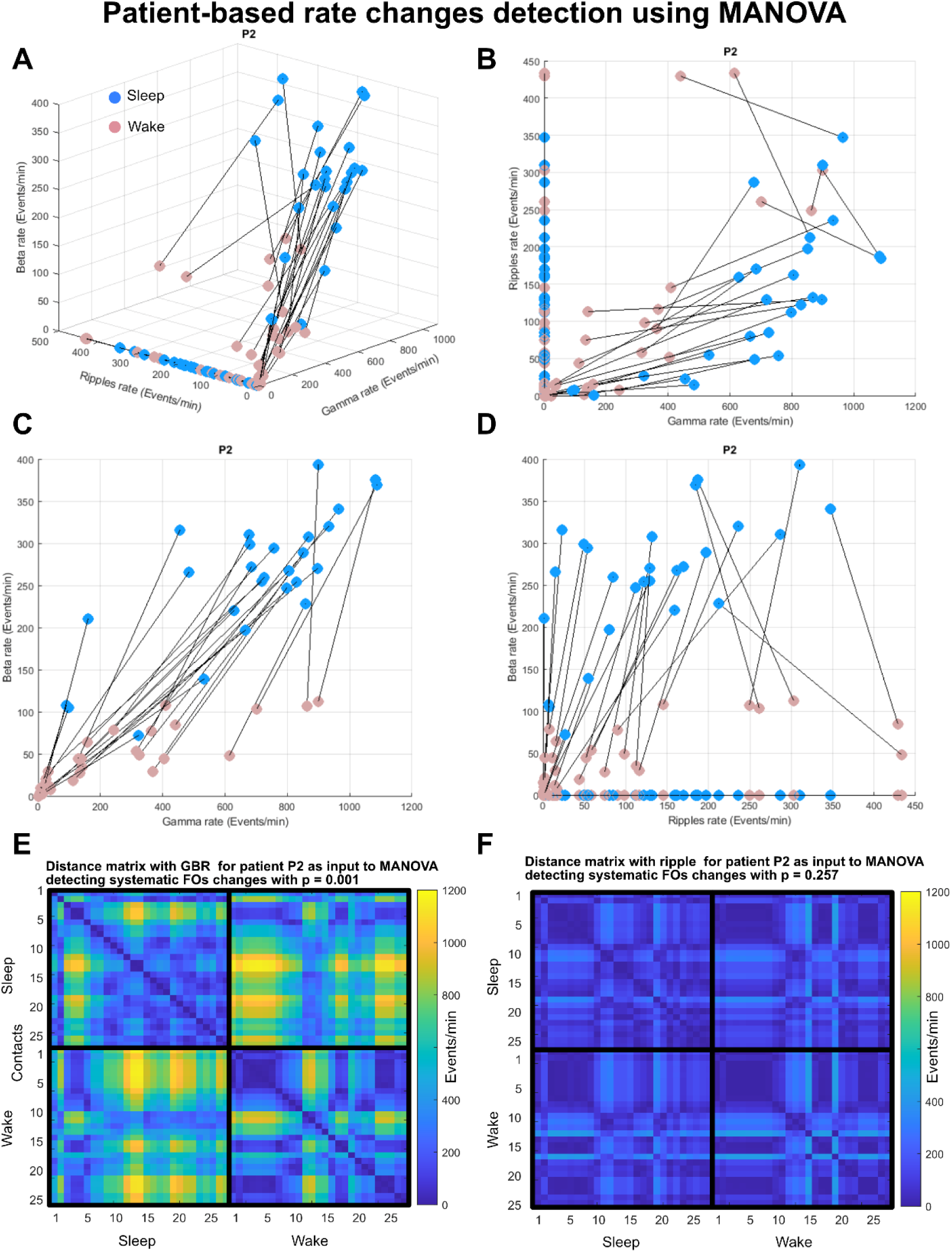
Changes in fast oscillation rates between wake and sleep across IZ contacts analyzed by MANOVA in patient P2. A: Representation of fast oscillation rates during wake (pink) and sleep (blue) in 3D space along beta, gamma and ripple axes. To compare the effectiveness of BGR versus ripple analysis alone, we replotted the same fast oscillation rate data points using only the ripple axis. Note that using all BGR provides larger separation between sleep and wake states. B: Shows 2D presentation of the 3D space presented in A with gamma rates on the x-axis and ripples on the y-axis. C and D: The same as B with different 2D perspectives. E: Distance matrix map of BGR fast oscillation rate differences measured between IZ contacts during the same states (upper left and lower right squares), or across wake and sleep states (lower left square). F: The same as in E but for ripples alone. Note that using BGR enables identification of differences between wake and sleep that were not distinguishable in this patient with ripple analysis alone.

### Statistical analysis

We used the Wilcoxon rank sum test (non-parametric) with FDR correction to account for multiple comparisons across the three frequency bands beta, gamma and ripple frequency bands. This testing was applied to test whether each band showed changes in FO rate at the population level for all patients (Figure 3).

The applied MANOVA explained above applies a non-parametric permutation-based statistical approach to test whether the F-value measured from the real data is larger than the ones produced by randomly shuffled data across the groups. If the real F-value is higher than 95% of the shuffled F-values then the effect is considered to be significant for a given patient.

We compared the outcome of MANOVA analysis regarding the detections of FO changes across wake and sleep comparisons for each frequency band (beta, gamma, ripple, and for all together BGR) in patient’s group perspectives: those with versus without sleep-related epilepsy, and those with frontal versus extra-frontal epilepsy.

We used chi-square tests to examine three clinical hypotheses, using a significance level of α < 0.05. We tested as well for associations between significant FO rate changes in:

1. Patients with versus without sleep-related epilepsy
2. Frontal versus non-frontal lobe epilepsy
3. FCD type II versus other histopathology (excluding three patients without histopathological FCD subtype classification)

## Results

### General FO rate changes between wake and sleep

At the group level, the raw FO rates in the beta, gamma, and ripple bands within the SOZ and the IZ showed a significant increase during sleep as compared to wake (p < 0.05; Wilcoxon rank sum test with FDR correction). Yet, these results might reflect a general effect of sleep on FO rates in all brain regions independent of whether these rates are due to SOZ or IZ. To test whether SOZ and IZ regions exhibit an increase in FO rates that is beyond a general effect, we adjusted SOZ and IZ rates by subtracting the rates measured from contacts of the group OTHER (Median raw values for groups, IZ, SOZ and OTHER see Suppl. 2). Because unadjusted values were not necessarily specific for the epileptic focus, they were not further evaluated, and we continued with the adjusted values.

### FO rates change in SOZ and IZ are beyond a general wake and sleep effect

At the group level and after adjustment for the general effect of sleep (details in methods), FOs in the SOZ still showed substantially higher rates during sleep as compared to wake for gamma oscillations and ripples (p < 0.01 and p < 0.05, respectively; Wilcoxon rank sum test with FDR correction). However, the effect seen before adjustment in beta oscillations did not survive after adjustment by the group OTHER (Figure 3A). FO rates in the IZ substantially increased from wake to sleep recordings for all three bands (p < 0.05, p < 0.01, p < 0.01; Wilcoxon rank sum test with FDR correction, for beta, gamma and ripple respectively, Figure 3B). These results demonstrate that the effects we report in SOZ and IZ are beyond any general effect that might be seen in other regions. Interestingly, a closer look at Figure 3A shows that the adjustment did not change the rates of ripples much in the SOZ in contrast with other bands that were affected by the adjustment procedure.

### Patient-based rate changes of beta, gamma, ripples and BGR

To apply a comprehensive analysis at the patient-by-patient level we used MANOVA, which provides a robust framework for analyzing FO rate changes across wake and sleep states, capturing the multidimensional complexity of BGR dynamics. This analysis considered the spatial organization of FO changes in feature space and allows simultaneous multiband analysis, ensuring a fair comparison between the combined BGR and the individual bands (see methods for more details). Using the MANOVA approach within the SOZ regions, significant increases were most frequent in the gamma band (10 patients), while BGR showed increases in seven patients (Figure 5A). Beta and ripple bands each showed significant increases in five patients. The numbers on the connecting lines indicate how many patients showed concordant findings between pairs of frequency bands, reflecting either concurrent increases or concurrent absence of significant changes.

**Figure 5:**
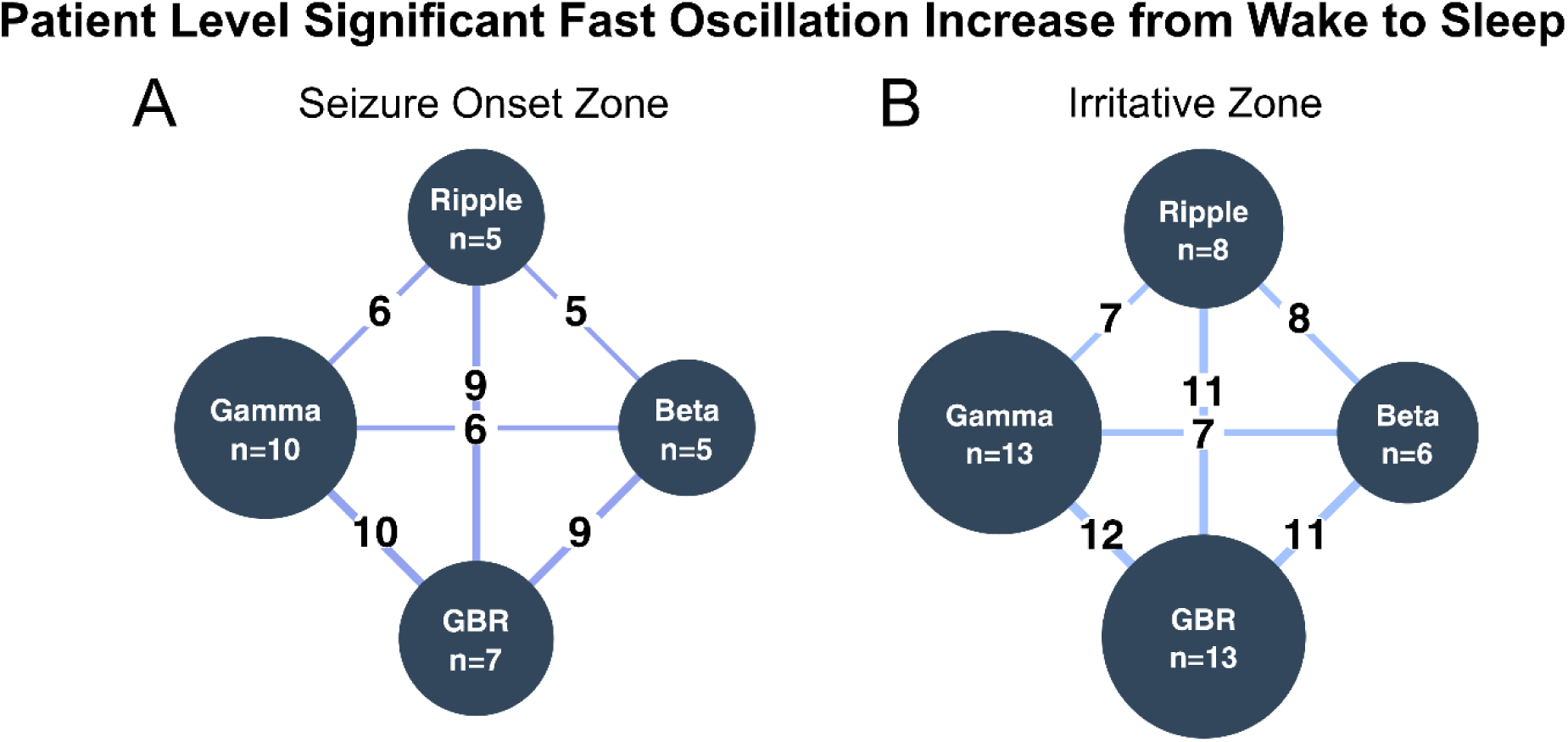
Multivariate Analysis of Variance (MANOVA) of FOs between wake and sleep. The MANOVA analysis was applied to different frequency bands to examine sleep-related changes in FO rates. Each dark blue circle represents the number of patients showing significant increases (p < 0.05) in the respective frequency band, with the size of circles proportional to the number of patients (n value shown for each band). Blue lines indicate concordant findings between pairs of frequency bands, with numbers showing how many patients had matching results between the connected bands. A: In the Seizure Onset Zone (SOZ), gamma oscillations showed the highest number of significant increases (n=10), followed by BGR (n=7), while beta and ripple bands each showed increases in five patients. The strongest concordance was observed between gamma and BGR (10 matching findings). B: In the Irritative Zone (IZ), both gamma and BGR showed frequent increases (n=13 each) with high concordance (12 matching findings). Ripple (n=8) and beta (n=6) bands showed more moderate increases, with varying degrees of concordance with other bands (7-11 matching findings). The varying patterns of concordance between different frequency bands suggest complementary contributions from each band in characterizing sleep-related changes.

Within the IZ, our MANOVA analysis revealed stronger and more consistent cross-frequency relationships compared to the SOZ (Figure 5B). The pattern between BGR and gamma band oscillations showed the highest concordance, with 12 patients showing agreement in their statistical findings. While beta and ripple oscillations also showed significant increases (ripple: eight patients, beta: six patients), they exhibited moderate levels of concordance with other frequency bands (7-11 matching findings between pairs).

### FOs in patients with sleep-related epilepsy, frontal lobe epilepsy and FCD type II

A comparison between patients with and without sleep-related epilepsy (defined as >60% of seizures occurring during sleep in presurgical video-EEG monitoring; details in Suppl. 1) revealed an association between sleep-related epilepsy and increased rates of gamma oscillations (p < 0.05, chi-square test, Bonferroni-corrected for four frequency bands: beta, gamma, ripple, broadband BGR). However, there was just a trend toward increased broadband BGR (p = 0.07, chi-square test, uncorrected) within the SOZ. No relevant associations were found for other frequency bands within the SOZ nor for any frequency bands within the IZ (Suppl. 2). There was also no relevant association between sleep-related FO-rate changes for frontal versus non-frontal SOZ or FCD type II versus other histopathological types (chi-square test, details Suppl. 2). No difference in sleep-related epilepsy was observed between patients with frontal and non-frontal epilepsy (p = 0.8, chi-square test, Suppl. 2).

## Discussion

### Information gained through broader bandwidth

Our analysis shows a systematic increase in FO rates from wake to sleep, specific to the epileptic focus, in iEEG recordings of patients with FCD. Within the SOZ, gamma oscillations showed the most substantial and consistent increases, with broadband oscillations (BGR) showing the second most frequent changes at the patient level. Significant changes in oscillation rates in the beta and ripple frequency bands were less common within the SOZ. Often, significant changes occurred simultaneously across multiple frequency bands, with gamma showing the highest concordance with other bands. It was less common to observe significant changes in beta and ripple bands in patients where gamma band and broadband analyses remained non-significant.

Within the IZ, the increase in FO rates from wake to sleep was more pronounced, with both gamma and broadband (BGR) frequencies showing equally strong and highly concordant changes. Beta and ripple frequency bands showed less frequently relevant changes.

These findings highlight that focusing on the gamma frequency band alone or including broadband frequencies yields the most distinct separation between wake and sleep. Beta and ripple frequency bands, while less effective for this distinction, often align with gamma and broadband BGR findings and provide additional relevant information within both SOZ and IZ in some patients. Gamma-band oscillations co-occurring with interictal epileptiform discharges in SOZ are known as good markers for the SOZ (Ren *et al*., 2015; Thomas *et al*., 2023). Similarly, it is known that increased rates of ripples are specific markers for the SOZ (Jacobs *et al*., 2010; Dimakopoulos *et al*., 2024). In our study, the specificity of ripples is reflected by the fact that they did not change much after adjustment by the group OTHER as they were rare outside the SOZ. However, we did not aim to evaluate the accuracy of those parameters in detecting the SOZ, but to characterize their changes between wake and sleep, which might help unraveling relevant differences between these brain states. Our current findings show that gamma oscillations perform best in identifying substantial increases in FO rates within the SOZ, while both gamma and broadband BGR excel in detecting changes in the IZ between wake and sleep iEEG recordings. The prominence of gamma oscillations within the SOZ might reflect more synchronized neuronal activity in the core epileptogenic zone, whereas the equal importance of BGR in the IZ could indicate more asynchronous broadband activity in the surrounding epileptogenic tissue. This aligns with our group’s previous observation of spectral peaks in the epileptic focus (Heers *et al*., 2018). The patient-specific variability in our current study suggests considering multiple frequency bands in the clinical analysis, with particular attention to the gamma frequency band.

A broad range of frequencies is known to be spatially related to the epileptic focus in patients with FCD. Beta and gamma oscillations contribute valuable information about the epileptic focus in FCD patients (Aubert *et al*., 2009). Gamma oscillations spatially overlap with dysplastic neurons in FCD type II, which establishes a very close pathophysiological relation (Rampp *et al*., 2021). Ripples are known to reflect disease severity in FCD patients (Kerber *et al*., 2014). Moreover, it has been demonstrated that gamma oscillations are not only increased within the SOZ but also in directly adjacent epileptogenic tissue, which might align with our finding that the inclusion of the irritative zone (IZ) improves detection of FO rate increases from wake to sleep (Sato *et al*., 2017). All these insights together reinforce the notion that a broadband approach could be relevant to capturing the complexity of the epileptic focus and the modulation of FOs between wake and sleep iEEG recordings of FCD patients.

### Sleep-related changes and potential mechanisms in FCD

Neuronal excitability in focal epilepsy undergoes significant changes during sleep. Studies using single-pulse electrical stimulation in patients with focal epilepsy show higher amplitude of early N1 responses during sleep, suggesting increased excitability followed by intense inhibition compared to wakefulness (Usami et al., 2015). In FCD patients, our observation of increased FO rates during sleep might reflect such heightened excitability, particularly as typical FCD discharge patterns (rhythmic spike, polyspike wave complexes at 1-3 Hz with fast discharges) not only become more frequent but also spread to surrounding non-lesional areas during NREM sleep (Tassi *et al*., 2012). Paradoxically, while increased excitability occurs during sleep, rates of interictal epileptiform discharges decrease during sleep in patients with FCD (Chassoux *et al*., 2012; Menezes Cordeiro *et al*., 2015).

The complex relationship between sleep and epileptic activity potentially stems from FCD’s integration within the thalamocortical network. Dysplastic neurons exhibit connectivity patterns resulting in stronger phase-amplitude coupling in regions with greater dysplastic neuron density (Rampp et al., 2021). This integration may explain our finding of significantly increased gamma oscillations specifically in patients with proven sleep-related epilepsy, with similar trends observed in broadband BGR. However, these broadband differences between wake and sleep are not unique to FCD patients, as similar patterns quantified through spectral slope changes have been observed in other types of focal epilepsy, suggesting increased excitation during sleep may be a broader phenomenon (Latreille *et al*., 2024). Contrary to previous reports (Khan *et al*., 2018), in our cohort we found no evidence that frontal lobe FCD patients experience more frequent sleep-related epilepsy compared to FCD patients with SOZ outside the frontal lobe. Whether sleep-related seizures are associated with lesion location or intrinsic properties of the dysplastic tissue remains undetermined, and the exact mechanisms by which sleep influences FCD epileptogenicity require further investigation (Menezes Cordeiro et al., 2015).

### Semi-automated detection of FOs

FO detection is crucial for epilepsy diagnostics and identifying the epileptic focus. Using semi-automated detection tools, like the Delphos toolbox (Roehri *et al*., 2018), we processed large datasets efficiently, allowing for consistent evaluation of FOs across beta, gamma, and ripple bands. The semi-automated detection enabled fast and standardized analyses of one-hour sleep and wake iEEG recordings, identifying frequency-specific changes that visual review might have missed. While this method reduces bias and speeds up analysis, it is not without limitations, such as susceptibility to filter artifacts and noise. To minimize errors, each recording was visually screened for artifacts, but the risk of including false positive detections remains, highlighting the need for further refinement of automated methods. Yet, such false positive detections are unlikely to be seen during one brain state but not another, which minimizes the bias of automated detection on our results.

Combining FOs from different frequency bands can be more informative than focusing on single frequency bands but is limited by the reviewer’s capacity to interpret large amounts of data. With the development and ongoing improvements of artificial intelligence, analyzing more complex information is becoming much easier and more available for clinical use, even for healthcare providers without coding experience (Faes et al., 2019).

### Limitations

In this study, we relied on semi-automated detections. The limitations of semi-automated detections are typically related to thresholding and might contain filter artifacts and technical artifacts (Thomschewski et al., 2020). However, the implemented detector efficiently avoided these artifacts by analyzing the data in the wavelet domain, and it was shown to be specifically insensitive to mistakenly identifying epileptic spikes as FOs (Roehri *et al*., 2016, 2018). The risk of erroneous marking was further reduced by visual inspection of each dataset.

Although the differentiation between wake and sleep recordings was done as thoroughly as possible, we did not perform sleep staging considering standard sleep stages.

### Future directions

In future studies, a more detailed differentiation of sleep stages and co-occurrences with K-complexes and sleep spindles will probably give even more insight (Frauscher and Gotman, 2019). Additionally, the extended analysis of sleep and wake intervals over multiple days could provide more valuable data (Alvarado-Rojas *et al*., 2014). Investigations of FO rate changes in relation to epileptic seizures, cross-frequency coupling, or data-driven analyses might also yield important findings (Le Van Quyen *et al*., 2005).

## Data Availability

The data used in this study contain confidential patient information and cannot be shared publicly. However, the data can be made available upon reasonable request from qualified researchers, subject to additional ethics board approval and data sharing agreements.

