## Supplementary material for "Sleep increases broadband fast oscillations in the epileptic focus of patients with focal cortical dysplasia": Suppl. 1

### Details iEEG, Histopathology and Postsurgical Outcome

IEEG was recorded using either subdural grid/strip electrodes or intracerebral depth EEG electrodes. Subdural EEG electrodes had a contact diameter of 4 mm with an exposed contact surface diameter of 2.3 mm and a center-to-center inter-contact distance of 10 mm (AD-tech, Racine, WI, USA). Intracerebral depth electrodes had a contact length of 2 mm with a center-to-center inter-contact distance of 4.5 mm and a contact diameter of 0.87 mm (AD-tech, Racine, WI, USA). In five patients, DIXI electrodes were used with a contact length of 2 mm, thickness of <0.8 mm, and 3.5 mm distance between contacts (Dixi-medical, Marchaux – Chaudefontaine, France).

Thirteen patients underwent stereo-EEG with multiple intracerebral depth electrodes, seven patients underwent subdural EEG, and two patients had a combination of both subdural and intracerebral depth. In 11 out of 22 patients, surface EEG was recorded simultaneously with iEEG.

After iEEG, all patients underwent epilepsy surgery. Histopathological specimens were classified according to the International League Against Epilepsy (ILAE) classification (Blümcke *et al.*, 2011). Histopathological examination revealed FCD type II in 12 patients, FCD type I in five patients, and mild malformation of cortical development (mMCD) in one patient. Four patients were categorized based on MRI criteria by an experienced neuroradiologist (H.U.) because the neuropathological samples were insufficient for accurate histopathological classification (Urbach *et al.*, 2023).

Favorable Engel class 1 postsurgical outcome was achieved in 15 of 22 (68%) patients. An unfavorable outcome occurred in 7 patients (Engel 2a: two patients, Engel 4b: five patients) (Engel, J. J. *et al.*, 1993) at least follow-up with a minimum follow-up of 12 months (details Table 1).

In accordance with the definition of sleep-related epilepsy, where the majority of seizures occur during sleep, we established the criterion that at least 60% of seizures should occur during sleep during presurgical video-EEG monitoring. Based on this criterion, 13 patients were identified as having sleep-related epilepsy. The ethics committee of the University Medical Center Freiburg approved this study, and all patients provided written informed consent for the scientific use of their clinical data recorded during their presurgical evaluation at the University Epilepsy Center Freiburg.

#### MRI analysis and identification of iEEG electrode positions

In our project, four high-resolution MRI sequences with isotropic voxels (edge length: 1 mm) were processed per patient. Three presurgical 3T MRI sequences (Trio or Prisma, Siemens, Erlangen, Germany) included a 3D T1 magnetization prepared (MPRAGE) sequence, a 3D fluid-attenuated inversion recovery (FLAIR) sequence, and, when available, a magnetization prepared rapid gradient echo (MP2RAGE) sequence with MRI morphometry (Demerath *et al.*, 2022) as part of the in house presurgical epilepsy MRI protocol (Urbach *et al.*, 2015). Additionally, all patients underwent a post-implantation 1.5T MRI (Avanto, Siemens, Erlangen, Germany) after iEEG electrode implantation.

All four MRI sequences were processed in the Matlab Toolbox (Matlab 2018b, Mathworks, Natick, MA, USA) Brainstorm (Tadel *et al.*, 2011). In Brainstorm, the positions of three landmarks (left and right preauricular points and nasion) were estimated using a 12-parameter affine transformation in SPM12 (Ashburner and Friston, 2005) aligned to the Montreal Neurological Institute ICBM 152 brain (Mazziotta *et al.*, 2001) without spatial normalization of individual MRIs. After rigid co-registration using SPM12, individual positions of iEEG electrode contacts were marked based on electrode positions, orientation, contact length, and inter-contact distance. The cortical surface was tessellated and assigned to the Automatic Anatomical Labeling atlas (Tzourio-Mazoyer *et al.*, 2002) using the Matlab toolbox CAT12 (Dahnke, Yotter and Gaser, 2013) as implemented in Brainstorm software. The FCD could be precisely visually identified in 15 patients (Urbach *et al.*, 2023).

Mazziotta, J., Toga, A., Evans, A., Fox, P., Lancaster, J., Zilles, K., Woods, R., Paus, T., Simpson, G., Pike, B., Holmes, C., Collins, L., Thompson, P., MacDonald, D., Iacoboni, M., Schormann, T., Amunts, K., Palomero-Gallagher, N., Geyer, S., Parsons, L., Narr, K., Kabani, N., Le Goualher, G., Feidler, J., Smith, K., Boomsma, D., Pol, H.H., Cannon, T., Kawashima,

R. and Mazoyer, B. (2001) ‘A Four-Dimensional Probabilistic Atlas of the Human Brain’, *Journal of the American Medical Informatics Association*, 8(5), pp. 401–430. Available at: https://doi.org/10.1136/jamia.2001.0080401.

Tadel, F., Baillet, S., Mosher, J.C., Pantazis, D. and Leahy, R.M. (2011) ‘Brainstorm: a user-friendly application for MEG/EEG analysis’, *Computational Intelligence and Neuroscience*, 2011, p. 879716. Available at: https://doi.org/10.1155/2011/879716.

Tzourio-Mazoyer, N., Landeau, B., Papathanassiou, D., Crivello, F., Etard, O., Delcroix, N., Mazoyer, B. and Joliot, M. (2002) ‘Automated Anatomical Labeling of Activations in SPM Using a Macroscopic Anatomical Parcellation of the MNI MRI Single-Subject Brain’, *NeuroImage*, 15(1), pp. 273–289. Available at: https://doi.org/10.1006/nimg.2001.0978.

Urbach, H., Mast, H., Egger, K. and Mader, I. (2015) ‘Presurgical MR Imaging in Epilepsy’, *Clinical Neuroradiology*, 25(2), pp. 151–155. Available at: https://doi.org/10.1007/s00062-015-0387-x.

Urbach, H., Scheiwe, C., Shah, M.J., Nakagawa, J.M., Heers, M., San Antonio-Arce, M.V., Altenmueller, D.-M., Schulze-Bonhage, A., Huppertz, H.-J., Demerath, T. and Doostkam, S. (2023) ‘Diagnostic Accuracy of Epilepsy-dedicated MRI with Post-processing’, *Clinical Neuroradiology*, 33(3), pp. 709–719. Available at: https://doi.org/10.1007/s00062-023-01265-3.
