## Supplementary material for "Sleep increases broadband fast oscillations in the epileptic focus of patients with focal cortical dysplasia": Suppl. 2

#### Median Detections per Frequency Band with Delphos Detector

| **Seizure Onset Zone (Events per minute: median Q1, Q3)** | | | | | |
|---|---|---|---|---|---|
| Beta | | Gamma | | Ripple | |
| Wake | Sleep | Wake | Sleep | Wake | Sleep |
| 27 (12, 63) | 73 (25, 105) | 70 (12, 295) | 73 (25, 105) | 192 (31, 422) | 230 (144, 862) |
| **Irritative Zone (Events per minute, median Q1, Q3)** | | | | | |
| Beta | | Gamma | | Ripple | |
| Wake | Sleep | Wake | Sleep | Wake | Sleep |
| 22 (11, 44) | 59 (26, 90) | 25( 12, 120) | 148 (86, 376) | 44 (12, 243) | 224 (130, 708) |
| **OTHER (Events per minute, median Q1, Q3)** | | | | | |
| Beta | | Gamma | | Ripple | |
| Wake | Sleep | Wake | Sleep | Wake | Sleep |
| 12 (8, 28) | 30 (11, 52) | 10 (3, 14) | 33 (20, 87) | 9 (2, 70) | 35 (10, 102) |

#### Numbers of intracranial EEG contacts per patient within different regions

| | mean | median | STD | range | total |
|---|---|---|---|---|---|
| SOZ | 14 | 11 | 12 | 1-45 | 299 |
| IZ | 29 | 29 | 16 | 3-52 | 628 |
| OTHER | 30 | 24 | 21 | 5-87 | 667 |

### Chi-Square Analyses:

### Analysis 1: Sleep-related epilepsy: distribution in frontal vs Non-frontal locations

SRE in frontal locations: 8/13 (61.5%)

SRE in Non-frontal locations: 5/9 (55.6%)

Chi-square = 0.079, p = 0.779

### Analysis 2: SRE vs oscillation patterns

| Seizure Onset Zone | Irritative Zone |
|---|---|
| Beta:<br>SRE: 4/13 significant (30.8%)<br>Non-SRE: 1/9 significant (11.1%)<br>Chi-square = 1.170, p = 0.279 | Beta:<br>SRE: 3/13 significant (23.1%)<br>Non-SRE: 4/9 significant (44.4%)<br>Chi-square = 1.119, p = 0.290 |
| Gamma:<br>SRE: 9/13 significant (69.2%)<br>Non-SRE: 1/9 significant (11.1%)<br>Chi-square = 7.246, **p = 0.007****<br>**(p = 0.04* Bonferroni-corrected)** | Gamma:<br>SRE: 10/13 significant (76.9%)<br>Non-SRE: 4/9 significant (44.4%)<br>Chi-square = 2.424, p = 0.119 |
| Ripple:<br>SRE: 4/13 significant (30.8%)<br>Non-SRE: 1/9 significant (11.1%)<br>Chi-square = 1.170, p = 0.279 | Ripple:<br>SRE: 6/13 significant (46.2%)<br>Non-SRE: 2/9 significant (22.2%)<br>Chi-square = 1.316, p = 0.251 |
| BGR:<br>SRE: 8/13 significant (61.5%)<br>Non-SRE: 2/9 significant (22.2%)<br>Chi-square = 3.316, p = 0.069 | BGR:<br>SRE: 9/13 significant (69.2%)<br>Non-SRE: 5/9 significant (55.6%)<br>Chi-square = 0.430, p = 0.51 |

### Analysis 3: Frontal vs Non-frontal oscillation patterns

| Seizure Onset Zone | Irritative Zone |
|---|---|
| Beta:<br>Frontal: 2/13 significant (15.4%)<br>Non-frontal: 3/9 significant (33.3%)<br>Chi-square = 0.976, p = 0.323 | Beta:<br>Frontal: 4/13 significant (30.8%)<br>Non-frontal: 3/9 significant (33.3%)<br>Chi-square = 0.016, p = 0.899 |
| Gamma:<br>Frontal: 5/13 significant (38.5%)<br>Non-frontal: 5/9 significant (55.6%)<br>Chi-square = 0.627, p = 0.429 | Gamma:<br>Frontal: 7/13 significant (53.8%)<br>Non-frontal: 7/9 significant (77.8%)<br>Chi-square = 1.316, p = 0.251 |
| Ripple:<br>Frontal: 3/13 significant (23.1%)<br>Non-frontal: 2/9 significant (22.2%)<br>Chi-square = 0.002, p = 0.962 | Ripple:<br>Frontal: 5/13 significant (38.5%)<br>Non-frontal: 3/9 significant (33.3%)<br>Chi-square = 0.060, p = 0.806 |
| BGR:<br>Frontal: 5/13 significant (38.5%)<br>Non-frontal: 5/9 significant (55.6%)<br>Chi-square = 0.627, p = 0.429 | BGR:<br>Frontal: 9/13 significant (69.2%)<br>Non-frontal: 5/9 significant (55.6%)<br>Chi-square = 0.430, p = 0.512 |

### Analysis 4: FCD Type II vs other pathologies

| **Seizure Onset Zone** | **Irritative Zone** |
|---|---|
| Beta:<br>FCD2: 3/12 significant (25.0%)<br>Other: 1/6 significant (16.7%)<br>Chi-square = 0.161, p = 0.688 | Beta:<br>FCD2: 3/12 significant (25.0%)<br>Other: 3/6 significant (50.0%)<br>Chi-square = 1.125, p = 0.289 |
| Gamma:<br>FCD2: 6/12 significant (50.0%)<br>Other: 3/6 significant (50.0%)<br>Chi-square = 0.000, p = 1.000 | Gamma:<br>FCD2: 8/12 significant (66.7%)<br>Other: 4/6 significant (66.7%)<br>Chi-square = 0.000, p = 1.000 |
| Ripple:<br>FCD2: 3/12 significant (25.0%)<br>Other: 2/6 significant (33.3%)<br>Chi-square = 0.138, p = 0.710 | Ripple:<br>FCD2: 3/12 significant (25.0%)<br>Other: 3/6 significant (50.0%)<br>Chi-square = 1.125, p = 0.289 |
| BGR:<br>FCD2: 5/12 significant (41.7%)<br>Other: 3/6 significant (50.0%)<br>Chi-square = 0.112, p = 0.737 | BGR:<br>FCD2: 6/12 significant (50.0%)<br>Other: 5/6 significant (83.3%)<br>Chi-square = 1.870, p = 0.171 |

FCD: focal cortical dysplasia, SRE: sleep related epilepsy, BGR: beta-gamma-ripple
